# Identifying and quantifying ESKAPEE pathogens in and around sinks in high burden hospitals

**DOI:** 10.64898/2026.03.13.26348341

**Authors:** Lindsay B. Saber, Melani Rojas, Ivory C Blakley, Shan Sun, Megan E.J. Lott, Anthony A Fodor, Carla Calderón Toledo, Joe Brown

## Abstract

Hospital-acquired infections driven by ESKAPEE pathogens (*Enterococcus faecium, Staphylococcus aureus, Klebsiella pneumoniae, Acinetobacter baumannii, Pseudomonas aeruginosa, Enterobacter spp*., *and Escherichia coli*) are highly prevalent. Premise plumbing, sinks and drains, seeds these organisms into patient environments via aerosolization and subsequent surface contamination. We measured viable ESKAPEE pathogens and overall microbial communities in and around sinks in two high-burden hospitals in La Paz, Bolivia, using culture and 16S rDNA sequencing. In a prospective observational study (May–August 2025), we collected 233 surface swabs and 39 air samples across four sink-related surface categories and in room air. Samples were plated on selective media for ESKAPEE identification and quantified as colony-forming units (CFU) normalized to 100 cm^2^ or 6000 L. DNA was extracted, and the full 16S rDNA gene was sequenced on PacBio Revio, analyzed via DADA2/QIIME2 and R. We detected viable presumptive ESKAPEE pathogens in 74.7% surface swabs and 74.4% air samples. Sink basins were most contaminated (mean 31CFU/100 cm^2^, 95 % CI16–46); concentrations declined with distance from the drain. *Klebsiella*/*Enterobacter* spp. showed the highest mean concentration across samples; *S. aureus* was most frequently detected (54.4% of samples). Hospital-specific differences were evident in culture positivity (Hospital A 85% vs. Hospital B 66.9%) and community composition (PERMANOVA P = 0.001; sample location explained 21.9% vs. 11.7% of variation). 16S profiling confirmed elevated relative abundances of *Klebsiella, Enterococcus*, and *Enterobacter* in basins relative to distant surfaces and air. The hospitals studied had high levels of ESKAPEE pathogens, underscoring the need for control measures.

## Introduction

Hospital Acquired Infections (HAIs) are a problem worldwide. Low- and Middle-Income Countries (LMICs) bear even more of the burden compared to High Income Countries(1). Antimicrobial Resistant (AMR) bacteria are a serious threat to public health, and many AMR infections are amplified in healthcare facilities. A specific group of bacteria, the ESKAPEE pathogens (*Enterococcus faecium, Staphylococcus aureus, Klebsiella pneumoniae, Acinetobacter baumannii, Pseudomonas aeruginosa, Enterobacter* spp., and *Escherichia coli*) (2) are the leading cause of HAIs, and are associated with high levels of multidrug resistance.

Premise plumbing, including sinks, basins, drains, and p-traps--the bend in a drain where water stands--are reservoirs for ESKAPEE pathogens (3). Bacteria are seeded in the p-trap via liquid disposal, and the conditions allow for multiplication (4). Sinks may present a risk of pathogen transmission to patients, as multiple outbreaks have been traced to bacteria originating in sinks (3, 5-19). Transmission from the drain is possible because when the faucet is turned on, the p-trap water is mechanically agitated and bacteria are aerosolized, creating the potential for aerosol transport that may result in inhalation or settling on surfaces and fomites (20).

The established sink transmission pathway presents a clear hazard in hospital settings, particularly those in LMICs, where ESKAPEE pathogens are abundant. Bolivia is a country in which there are many AMR HAIs, but the source of these infections is often unclear due to limited surveillance and diagnostic capacity (21). Therefore, we sought to identify and quantify the ESKAPEE pathogens in and around sinks in two Bolivian hospitals. We hypothesized that ESKAPEE pathogens would be variably present in and around hospital sinks, detected by both culture and 16S methods.

## Methods

### Setting and Design

We conducted a prospective observational study of ESKAPEE pathogens in and around sinks in two hospitals in the metropolitan area of La Paz, Bolivia, where HAIs are highly prevalent (21, 22). Hospital A is smaller with a higher density, while Hospital B is larger, with more space for separate patient care. Most of the sampled sinks were in common rooms and mostly used by medical and cleaning staff. Our primary objective was to identify and quantify the ESKAPEE pathogen presence in and around hospital sinks, with some additional environmental samples collected opportunistically. These samples were collected as a baseline for a disinfection intervention in the two hospitals(23).

### Sample Collection

#### Sample Collection and Processing

Within each hospital, we collected environmental samples from patient rooms, preparation rooms, cleaning rooms, and reception spaces. We grouped the swab sample locations in four distinct categories: (1) basin interior, the inside of the sink basin, (2) sink surfaces, the remaining surfaces of the sink excluding the basin interior (e.g. sink handles and sink edge), (3) near-sink surfaces, the surfaces less than 1m from the basin, and (4) high touch surfaces, which were located more than 1m away from the sink and which the hospital staff indicated is higher risk for pathogen transmission due to high contact. The area of each sample varied but was measured and recorded. We collected air samples from each room.

#### Surface Samples

We swabbed each surface of interest with an Isohelix Swabs (Cat No. SK-3S), pre-wet with sterile Dey-Engley (DE) Neutralizing Buffer (Cat No. D3435). Following sampling, we eluted each swab into 1 mL of sterile DE Buffer and transported at 4°C until sample processing. Upon receipt in the laboratory, we vortexed swabs for 30 seconds. From the eluate, we plated 100 µL onto duplicate agar plates of CHROMagar Orientation (Cat No. RT412), CHROMagar ESBL (Cat No. ESRT2), CHROMagar Acinetobacter (Cat No. AC092), and CHROMagar Pseudomonas (Cat No. PS832). We incubated all agar media at 37°C for 24 hours, except CHROMagar Pseudomonas at 30°C. From the remaining eluate we stored 500 µL in 2X DNA/RNA Shield (Zymo, Cat No. R1200).

#### Air Samples

We collected air samples with the AirPrep Cub Sampler (Innovaprep, Drexel, MO) for 30 min at 200 L min^-1^. We transported cassettes at 4°C until sample processing. Upon receipt in the laboratory, we eluted bioaerosols from the cassette into 6 mL of InnovaPrep FluidPrep EasyElute Elution Buffer (Cat No. HC08018-T). We added equal parts DE Neutralizing Buffer, spun down to concentrate, and eluted the supernatant for a final volume of 1mL. We plated 100 µL of the concentrated sample onto duplicate agar plates of CHROMagar Orientation, CHROMagar ESBL, CHROMagar Acinetobacter, and CHROMagar Pseudomonas as described above. We incubated all agar media at 37°C for 24 hr, except CHROMagar Pseudomonas at 30°C. From the remaining eluate, we stored 500µL in 2X DNA/RNA Shield.

### Culture Analysis

#### Detection of ESKAPEE Pathogens by Culture

Following incubation, we examined agar plates for *Acinetobacter* spp., *E. coli, Enterococcus* spp., *Klebsiella/ Enterobacter* spp., *P aeruginosa*, and *S. aureus* based on colony color and morphology, as stated in manufacturer guidelines. We recorded the CFUs and presence or absence of presumptive ESKAPEE pathogen for each swab and air sample analyzed. The morphology of *Klebsiella* spp., *Enterobacter* spp. and *Citrobacter* spp. are identical on CHROMagar Orientation and ESBL media. Therefore, any reference of *Klebsiella*/ *Enterobacter* spp. via culture could presumptively be any one of the three genera.

Analysis was completed in R (v4.4.1). All surface samples were normalized to 100cm^2^ while all air samples are analyzed at the collected 6000 LPM. All means are arithmetic. Positivity rates are a simple percentage of positive samples in each category.

### Molecular Analyses

#### Nucleic Acid Isolation

We transported eluates preserved in DNA/RNA shield at ambient temperature to the laboratory at the University of North Carolina at Chapel Hill. We isolated DNA from the samples using the ZymoBIOMICS DNA/RNA Mini Prep Kit (Zymo, Cat No. R2002).

#### 16S Library Preparation and Sequencing

We submitted isolated nucleic acids to the Duke Microbiome Core Facility for library preparation and long-read sequencing of the full 16s rDNA gene on PacBio. Total DNA extracted from each sample was quantified via Qubit and normalized to 1ngµl^-1^ for each sample. 2ng of total DNA was subject to PCR amplification of the full-length 16S rRNA gene as outlined in the PacBio Kinnex 16S kit (PacBio, Cat No. 103-072-100), using Phusion Plus PCR Master Mix (Thermoscientific, Cat No. F631L), and 27F-1492R universal primer set (5’-AGRGTTYGATYMTGGCTCAG-3’ and 5’-RGYTACCTTGTTACGACTT-3’, respectively) containing unique barcodes and Kinnex adaptors at a final concentration of 0.3uM. The cycling conditions were: 30 seconds at 98°C; 30 cycles of denaturation at 98°C for 10 seconds, annealing at 57°C for 20 seconds, and extension at 72°C for 75 seconds; a final extension at 72°C for 5 minutes.

Completed PCR reactions were visualized on an E-Gel (Invitrogen) to ensure that amplicon size was correct (∼1500bp), and that each sample amplified appropriately. Amplicon libraries were ubsequently pooled at different volumes based on gel band intensity (0.5, 1.25, 2.5, 5, or 10µl per reaction). Each library pool was cleaned and concentrated using 1.1x volume of SMRTbell® cleanup beads (PacBio, Cat No. 103-306-300) and eluted in 100µl of Low TE Elution Buffer (PacBio, Cat No. 102-178-400). Cleaned libraries were stored at -20°C prior to Kinnex PCR for concatenation and circularization, performed according to the PacBio Kinnex 16S kit’s published protocol with no modifications. Size selected and cleaned libraries were loaded onto a PacBio SMRT® Cell and sequenced on the Revio system (PacBio) in the Duke Sequencing and Genomic Technologies Shared Resource.

#### 16S Analysis

We analyzed demultiplexed reads of the 16S rRNA amplicon sequencing with DADA2 and QIIME2 following the developers’ instructions (24). We performed denoising, dereplication, ASV inference, and chimera removal with DADA2 PacBio long reads pipeline(25). We assigned taxonomy with QIIME2 feature classifier using the GTDB database (release 220). We normalized taxonomic abundance to correct for different sequencing depth as previously described (26). We performed statistical analyses with R (4.3.1). We computed beta-diversity using Bray-Curtis dissimilarity and visualized with principal coordinates analysis (PCoA) using R function ‘capscale’ in the package ‘vegan’. We analyzed the differences in microbial community profiles between groups with PERMANOVA test using function ‘adonis2’ in the same package with 999 permutations. We used the Shannon index for analyzing alpha-diversity. We analyzed differential abundance of individual taxa between groups using non-parametric Wilcoxon rank sum test. We corrected p-values for multiple hypothesis testing using the Benjamini-Hochberg method.

## Results

### Microbial Culture

Between May 2025 and August 2025, we collected a total of 272 samples in two hospitals during 12 separate sampling episodes. Of these samples, 39 were air samples, 233 were surface swabs. Surface swab samples were one of four locations: sink basin (n=37), sink surfaces (n=89), near-sink surfaces (n=39), and high touch (n=68).

Overall, 74.7% (174/233) of swab and 74.4% (29/39) of air samples were positive for at least one viable, presumptive ESKAPEE pathogen. Basin interior samples were the most contaminated across both hospitals, with an overall mean concentration of 31 CFU/100 cm^2^, 95% CI: 16-46. The least contaminated samples were air samples (mean concentration of 1 CFU/100 cm^2^, 95% CI: 0-2). *S. aureus* was present in most samples (54.8%, 149/272), but *Klebsiella*/ *Enterobacter* spp. was the most concentrated when analyzed across all samples (mean 100 CFU/100 cm^2^, 95% CI: 60-200). Table 1 reports the concentration of the ESKAPEE pathogens in each sample type, per hospital. Table 2 reports the binary outcome of presence or absence of ESKAPEE pathogens.

**Table 1.** Arithmetic Man (95%CI) of ESKAPEE pathogen Colony Forming Units (CFUs) by hospital and sample type.

| Mean (95% CI) for each presumptive ESKAPEE pathogen by sample type and hospital |  |  |  |  |  |  |  |  |  |  |  |
| --- | --- | --- | --- | --- | --- | --- | --- | --- | --- | --- | --- |
| Surfaces: per 100cm<sup>2</sup>, Air: per 6,000 liters |  |  |  |  |  |  |  |  |  |  |  |
|  | Hospital A |  |  |  |  | Hospital B |  |  |  |  | All Samples |
|  | Basin Interior | Sink Surfaces | Near- Sink Surfaces | High Touch | Air | Basin Interior | Sink Surfaces | Near- Sink Surfaces | High Touch | Air |  |
| Acinetobacter | 133 (84, 182) | 173 (135, 211) | 0 (0, 0) | 0 (0, 0) | 0 (0, 0) | 25 (0, 58) | 177 (158, 195) | 0 (0, 0) | 1 (0, 1) | 0 (0, 0) | 70 (30, 100) |
| E. coli | 18 (9, 28) | 4 (4, 5) | 72 (49, 95) | 0 (0, 0) | 0 (0, 0) | 2 (0, 5) | 3 (3, 3) | 0 (0, 0) | 3 (2, 4) | 0 (0, 1) | 10 (0, 20) |
| Enterococcus | 44 (27, 60) | 35 (10, 59) | 0 (0, 1) | 0 (0, 0) | 0 (0, 0) | 0 (0, 0) | 1 (1, 1) | 0 (0, 0) | 0 (0, 0) | 0 (0, 0) | 6 (0, 10) |
| Klebsiella/ Enterobacter | 435 (369, 502) | 266 (221, 310) | 73 (47, 99) | 54 (36, 72) | 1 (0, 3) | 57 (7, 106) | 138 (122, 154) | 32 (30, 34) | 6 (3, 8) | 1 (0, 3) | 100 (60, 200) |
| Pseudomonas | 264 (206, 322) | 65 (30, 99) | 0 (0, 0) | 16 (10, 22) | 0 (0, 0) | 15 (0, 30) | 227 (205, 249) | 0 (0, 0) | 2 (0, 3) | 0 (0, 0) | 80 (40, 100) |
| S. aureus | 93 (58, 128) | 68 (59, 77) | 151 (137, 165) | 50 (43, 58) | 7 (1, 14) | 39 (11, 67) | 164 (150, 178) | 59 (50, 68) | 84 (75, 93) | 6 (2, 9) | 90 (60, 100) |
| Any ESKAPEE pathogen | 165 (0, 348) | 102 (0, 230) | 49 (0, 132) | 20 (0, 65) | 1 (0, 4) | 23 (0, 66) | 118 (0, 236) | 15 (0, 50) | 16 (0, 38) | 1 (0, 3) | NA |

**Table 2.** Positivity rates for presumptive ESKAPEE pathogens amongst each sample type for each hospital.

| Presumptive viable ESKAPEE pathogens by hospital and sample type |  |  |  |  |  |  |  |  |  |  |  |
| --- | --- | --- | --- | --- | --- | --- | --- | --- | --- | --- | --- |
| Percent of samples positive (% (n/N)) |  |  |  |  |  |  |  |  |  |  |  |
|  | Hospital A |  |  |  |  | Hospital B |  |  |  |  | All Samples |
|  | Basin Interior | Sink Surface | Near Sink | High Touch | Air | Basin Interior | Sink Surface | Near Sink | High Touch | Air |  |
| Acinetobacter | 27.8% (5/18) | 36% (9/25) | 4% (1/25) | 0% (0/32) | 0% (0/19) | 21.1% (4/19) | 20.3% (13/64) | 0% (0/14) | 5.6% (2/36) | 0% (0/20) | 12.5% (34/272) |
| E. coli | 22.2% (4/18) | 20% (5/25) | 8% (2/25) | 0% (0/32) | 5.3% (1/19) | 10.5% (2/19) | 7.8% (5/64) | 7.1% (1/14) | 22.2% (8/36) | 5% (1/20) | 10.7% (29/272) |
| Enterococcus | 22.2% (4/18) | 4% (1/25) | 4% (1/25) | 0% (0/32) | 0% (0/19) | 0% (0/19) | 3.1% (2/64) | 0% (0/14) | 2.8% (1/36) | 0% (0/20) | 3.3% (9/272) |
| Klebsiella/Enterobacter | 72.2% (13/18) | 68% (17/25) | 36% (9/25) | 25% (8/32) | 31.6% (6/19) | 31.6% (6/19) | 34.4% (22/64) | 21.4% (3/14) | 30.6% (11/36) | 45% (9/20) | 38.2% (104/272) |
| Pseudomonas | 55.6% (10/18) | 32% (8/25) | 0% (0/25) | 3.1% (1/32) | 0% (0/19) | 26.3% (5/19) | 20.3% (13/64) | 0% (0/14) | 5.6% (2/36) | 0% (0/20) | 14.3% (39/272) |
| S. aureus | 22.2% (4/18) | 44% (11/25) | 76% (19/25) | 68.8% (22/32) | 68.4% (13/19) | 26.3% (5/19) | 35.9% (23/64) | 64.3% (9/14) | 77.8% (28/36) | 70% (14/20) | 54.4% (148/272) |
| Any ESKAPEE pathogen | 83.3% (15/18) | 92% (23/25) | 92% (23/25) | 75% (24/32) | 73.7% (14/19) | 52.6% (10/19) | 60.9% (39/64) | 64.3% (9/14) | 83.3% (30/36) | 75% (15/20) | 74.3% (202/272) |

Of all swab samples taken, Hospital A swabs were 85% (85/100) and Hospital B samples were 66.9% (89/133) positive via culture methods. Air sample positivity was more comparable; Hospital A 73.7% (14/19) and Hospital B 75% (15/20) positive via culture methods. In Hospital A, basin interior samples had the highest quantity of ESKAPEE pathogens, mean 165 CFU/100cm^2^, 95% CI: 0-348. However, sink surface and near-sink samples had higher rates of ESKAPEE pathogen detection, both 92% (23/25). In Hospital B, sink surface samples contained the most culturable ESKAPEE pathogens (mean 118 CFU/100cm^2^, 95% CI 0-236), but high touch samples (83.3%, 30/36) were the most frequently contaminated.

### 16S Amplicon Sequencing

#### Microbial community compositions of surface sample

We used full length sequencing of 16S rRNA gene (PacBio) to characterize the microbial communities of the surface and air samples.

In Hospital A, the PCoA plots indicated that sink surface and basin interior samples clustered separately and were distinct from the other sample types (Fig. 1A). By contrast, at Hospital B, the clusters of sink surface, basin interior and high touch samples are overlapping with each other but distinct from near-sink surfaces and air samples, indicating that these samples were less differentiated by surface type around the sink (Fig. 1B). PERMANOVA tests indicated that the microbial compositions were significantly associated with sample locations (P=0.001) in both hospitals, but locations explained 21.9% variation in Hospital A samples and 11.7% in Hospital B samples.

**Figure 1.**
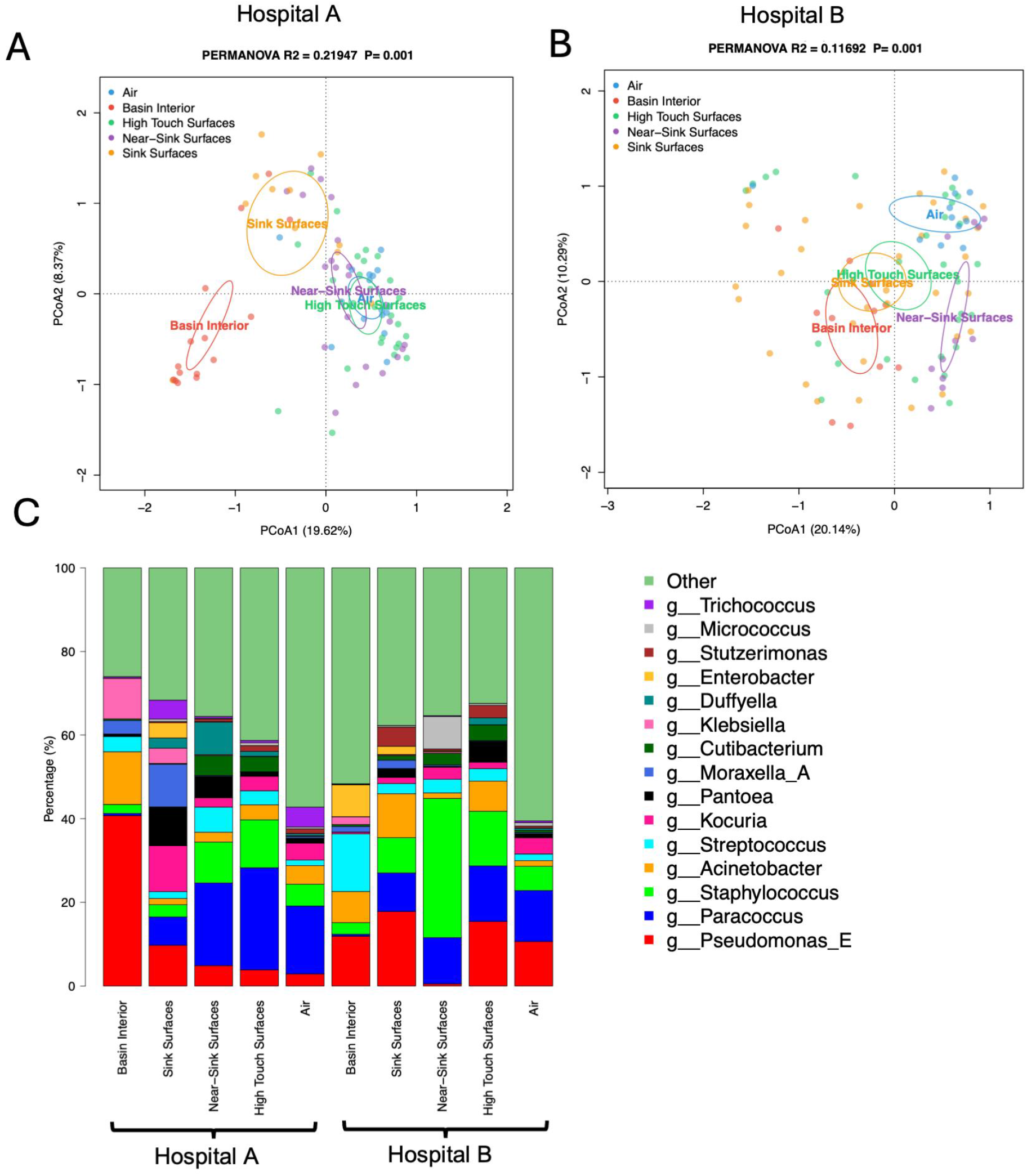
Principal Coordinates Analysis (PCoA) and average microbial composition of surface and air samples. (A) and (B) PCoA plots of Bray-Curtis dissimilarity at the genus level colored by sample type in Hospital A and Hospital B. Eclipses indicate 95% confidence limits. (C) The bar plot illustrates the average genus-level taxonomic composition of microbial communities grouped by surface category and air.

A bar plot of genus-level average taxonomic composition highlighted differences between sample types and between hospitals (Fig. 1C). Hospital A basin interior samples had a higher proportion of *Pseudomonas* spp., whereas Hospital B basins showed more *Streptococcus* spp. and near-sink surfaces showed more *Staphylococcus* spp.

#### Presence of ESKAPEE Pathogens in 16S data

In examining the ESKAPEE pathogens across the surface and air samples, we found differential abundance depending on the location and the hospital (Fig. 2). Sink surface and basin interior samples generally harbored higher levels of *Pseudomonas* spp., *Klebsiella* spp., and *Enterobacter* spp. compared to other surfaces and air samples, but they showed lower levels of *Staphylococcus* spp. Differences were also evident between the two hospitals: the Hospital B had a greater abundance of *Acinetobacter baumannii Staphylococcus* spp., whereas Hospital A showed higher levels of *Pseudomonas* spp.

**Figure 2.**
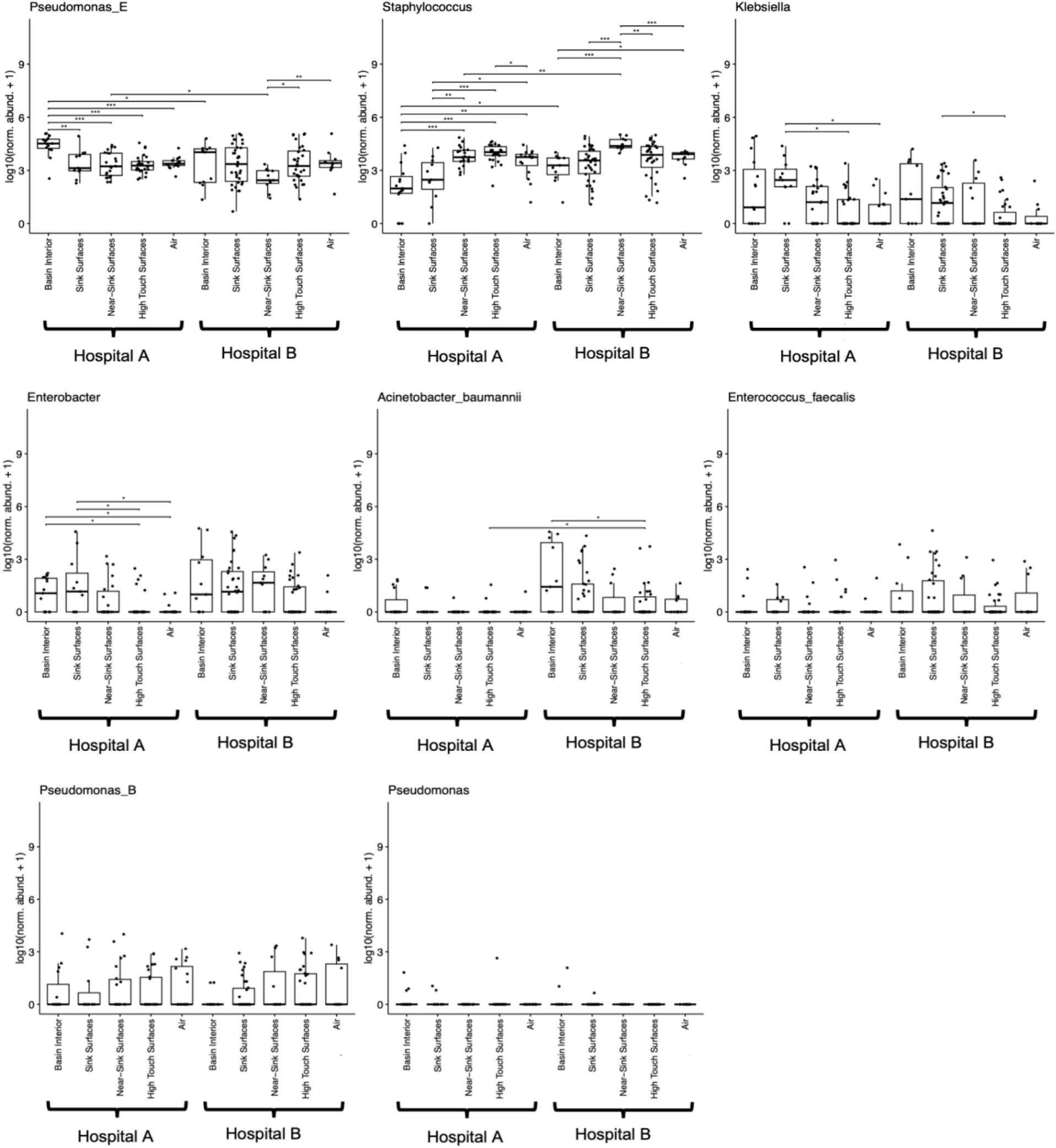
ESKAPEE pathogen abundance across sample types. Species level was used when classified, otherwise genus level abundance was visualized. Statistical analyses were performed with pairwise Wilcoxon rank-sum tests, restricted to same-hospital or same-sample-type pairs. P-values were corrected using the Benjamini-Hochberg method to correct for multiple comparisons (*** FDR < 0.001, ** FDR < 0.01, * FDR < 0.1).

## Discussion

In this study, we have described an integrated approach to identifying and quantifying microbial communities in a high-transmission setting, highlighting the potential utility of applying both culture and sequencing methods to more fully characterize microbial hazards. The presence and viability of ESKAPEE pathogens in and around sink areas suggests potential for transmission to patients and staff occupying these spaces. The hospitals in this study exhibit higher prevalences of ESKAPEE pathogens in and around sinks in comparison to other similar studies, even when resistant infections are known(9, 11, 12, 15, 17, 27).

We found viable presumptive ESKAPEE pathogens in all sample types across both hospitals. Basin interior samples were the most contaminated. As the swab sample categories increased in distance from the sink, the concentration of culturable ESKAPEE pathogens generally decreased. This trend is consistent with sink drains being a reservoir for pathogen growth(5, 28, 29). *Klebsiella*/ *Enterobacter* spp. was the most frequently isolated ESKAPEE pathogen and was found across all sample types.

The similarity between the 16S sequencing results differed between the hospitals. PCoA plots showed the basin interior and sink surfaces to be significantly different from the other samples in Hospital A, whereas Hospital B showed these clusters overlapping. The high relative abundance of *Klebsiella* spp. in the 16S results aligned with the high presence in the culture data. However, the low levels of *Staphylococcus* spp. in the 16S data conflict with the high levels of presumptive *S. aureus* in the culture data. These discrepancies are consistent with other studies, which have observed significant differences in bacterial detection when using both methods (30, 31).

Our data suggests the prevalence of ESKAPEE pathogens is different between the hospitals. The two settings differ in economic resources, which may affect the availability of cleaning staff, maintenance, and other factors that could contribute to effective control measures that reduce HAIs or environmental transport of pathogens generally. A dedicated cleaning team is present in Hospital B, while the responsibility to disinfect is on nurses in Hospital A. Previous studies have specified that environmental disinfection schedules, techniques, and distinct cleaning staff significantly impact pathogen load in environmental samples(6, 32, 33). Alternatively, the differences in microbial and molecular profiles may be attributed to the sink material or source water. Hospital A had more porcelain sinks, while Hospital B had more stainless steel; a wide range of bacterial survivorship has been reported on both materials (34, 35), potentially contributing to the differences in sink microbiomes. Factors such as distance-from-source, water temperature, and water aeration can significantly affect the water microbial concentrations (36).

Country-level data on the burden of AMR HAIs in Bolivia is limited. One study examined hospital acquired blood infections in Bolivia found *Klebsiella pneumoniae* to be the most frequently isolated bacteria, and *Enterobacter* spp. the third most isolated (21). *Enterococcus* spp. was also on the list of most isolated bacterium. This is consistent with our findings, as *Kelbsiella/ Enterobacter* spp. were both found in the highest concentrations when analyzed via culture. This pattern highlights a consistency of viable bacteria in the environment and those that are confirmed to cause HAIs, confirming a higher risk of patient infection per established models (37). As the sink-to-patient transmission pathway is well established, our results present a higher risk of ESKAPEE pathogen infection when compared to hospitals in other regions, such as the United States (37-42).

Our work underscores the need for developing novel solutions to control ESKAPEE pathogen transmission in this and similar settings. A range of mitigation strategies for limiting plumbing-associated infection transmission have been attempted, with variable success (43-45). These include chemical disinfectant treatments (44, 46, 47), mechanical cleaning (27), vibration and heat (43) or heat combined with disinfectant (48), bacteriophage treatment (49), and other measures, though recolonization of sinks and even horizontal transmission between sinks and drains in a plumbing network (50, 51) has been shown to occur rapidly and repeated treatments are usually necessary once a drain network has been colonized. Other well-characterized methods for reducing biofilm-associated risks – like the addition of bacteriostatic or biocidal metals (copper, silver, selenium compounds) (52), quorum sensing (53), and probiotic treatment (e.g., with *Bacillus*) (54) have been proposed but have not been widely evaluated. Sink replacement, drain replacement, and point of use filtration have been proposed but are not considered effective long-term strategies; maintaining sterility is also not feasible (55). New facilities can be colonized quickly (27) and quick response to known colonization may be key (27) in excluding pathogens from drain networks, but repeated cleaning is usually needed and even that may not be effective over time (45).

### Limitations

These results should be interpreted alongside the limitations of our sample collection methods and analysis. Air samples were the least contaminated, which may be a result of limited aerosolized pathogens. However, the AirPrep sampler is known to be more suited for sequencing-based analysis than culture-based detection, and the viability of pathogens may have been compromised while using this method. Sample collection was not coordinated with cleaning schedules, which may have affected bacterial presence. The culture and 16S resolution are not consistently granular to the species level, only the genus level. More information on the species and antimicrobial resistance gene presence could be provided through molecular characterization. Specifically for *Acinetobacter* spp., *Enterococcus* spp., *Klebsiella/ Enterobacter* spp., and *Pseudomonas* spp., the culture methods can only identify bacteria to the genus level. For these reasons, we suspect our samples contain common, non-pathogenic bacteria which belong to the ESKAPEE genus classifications, in addition to true ESKAPEE pathogens.

## Conclusion

We report the prevalence and concentration of ESKAPEE pathogens in environmental samples from two hospitals in Bolivia. Both culture-based and molecular methods demonstrate the sinks harbor epidemiologically important pathogens, such as *Klebsiella, Acinetobacter, Enterococcus, Pseudomonas*, and *S. aureus*. Considering the growing threat of AMR HAIs in Bolivia and globally, our findings highlight the need to mitigate the transmission of ESKAPEE pathogens from sinks, surfaces, and air to vulnerable patients.

## Data Availability

All data produced in the present study are available upon reasonable request to the authors

## Acknowledgements

The authors appreciate the hospital administrators, doctors, nurses, and cleaning staff for cooperation and access to sampling sites. This work was supported primarily by the Engineering Research Centers Program of the National Science Foundation under NSF Cooperative Agreement No. EEC-2133504. This research was funded in part by a grant from the NIH (T32ES007018).

